# Using rapid online surveys to assess perceptions during infectious disease outbreaks: a cross-sectional survey on Covid-19 among the general public in the United States and United Kingdom

**DOI:** 10.1101/2020.03.13.20035568

**Authors:** Pascal Geldsetzer

**Author notes:** Corresponding author: Pascal Geldsetzer MD PhD MPH, Division of Primary Care and Population Health, Department of Medicine, Stanford University, 1265 Welch Road, Stanford, CA 94305, USA.

## Abstract

**Background:** Given the extensive time needed to conduct a nationally representative household survey and the commonly low response rate in phone surveys, rapid online surveys may be a promising method to assess and track knowledge and perceptions among the general public during fast-moving infectious disease outbreaks.

**Objective:** To apply rapid online surveying to determine knowledge and perceptions of coronavirus disease 2019 (Covid-19) among the general public in the United States (US) and the United Kingdom (UK).

**Methods:** An online questionnaire was administered to 3,000 adults residing in the US and 3,000 adults residing in the UK who had registered with Prolific Academic to participate in online research. Strata by age (18 - 27, 28 - 37, 38 - 47, 48 - 57, or ≥58 years), sex (male or female), and ethnicity (White, Black or African American, Asian or Asian Indian, Mixed, or “Other”), and all permutations of these strata, were established. The number of participants who could enrol in each of these strata was calculated to reflect the distribution in the US and UK general population. Enrolment into the survey within the strata was on a first-come, first-served basis. Participants completed the questionnaire between February 23 and March 2 2020.

**Results:** 2,986 and 2,988 adults residing in the US and the UK, respectively, completed the questionnaire. 64.4% (1,924/2,986) of US and 51.5% (1,540/2,988) of UK participants had a tertiary education degree. 67.5% (2,015/2,986) of US participants had a total household income between $20,000 and $99,999, and 74.4% (2,223/2,988) of UK participants had a total household income between £15,000 and £74,999. US and UK participants’ median estimate for the probability of a fatal disease course among those infected with SARS-CoV-2 was 5.0% (IQR: 2.0% – 15.0%) and 3.0% (IQR: 2.0% – 10.0%), respectively. Participants generally had good knowledge of the main mode of disease transmission and common symptoms of Covid-19. However, a substantial proportion of participants had misconceptions about how to prevent an infection and the recommended care-seeking behavior. For instance, 37.8% (95% CI: 36.1% – 39.6%) of US and 29.7% (95% CI: 28.1% – 31.4%) of UK participants thought that wearing a common surgical mask was ‘highly effective’ in protecting them from acquiring Covid-19. 25.6% (95% CI: 24.1% – 27.2%) of US and 29.6% (95% CI: 28.0% – 31.3%) of UK participants thought it prudent to refrain from eating at Chinese restaurants. Around half (53.8% [95% CI: 52.1% – 55.6%] of US and 39.1% [95% CI: 37.4% –40.9%] of UK participants) thought that children were at an especially high risk of death when infected with SARS-CoV-2.

**Conclusions:** The distribution of participants by total household income and education followed approximately that of the general population. The findings from this online survey could guide information campaigns by public health authorities, clinicians, and the media. More broadly, rapid online surveys could be an important tool in tracking the public’s knowledge and misperceptions during rapidly moving infectious disease outbreaks.

## Introduction

When faced with rapidly moving infectious disease outbreaks, as is the case with coronavirus disease 2019 (Covid-19), assessing knowledge and perceptions of relevant populations has to be accomplished in a short time frame if the findings are to be informative to the public health response. Population-representative household surveys generally take many months of preparation and data collection.[1] Phone surveys are faster to conduct but are increasingly suffering from low response rates (typically well below 10%[2]),[2] which can be a major source of bias even when extensive weighting adjustments are undertaken.[3] In addition, unless they use interactive voice response (which tends to further decrease the response rate [4]), phone surveys require substantial human resources to conduct the interviews. Given these limitations, rapid online surveys, which demand minimal human resources (beyond those needed to design the questionnaire) and could reach large numbers of respondents in a short time frame, may be a valuable tool to assess (and monitor over time) knowledge and perceptions of an infectious disease in the midst of an outbreak.

Covid-19 was first reported in Wuhan, China, in December 2019.[5] On March 11^th^ 2020, the World Health Organization (WHO) declared Covid-19 a pandemic;[6] and by March 17^th^ 2020, there were more than 200,000 confirmed cases and over 8,000 reported deaths from Covid-19 across the globe.[7] The course of the Covid-19 epidemic in the United States (US) and the United Kingdom (UK) will likely be strongly impacted by how the population behaves, which in turn is influenced by what people know and believe about this disease.[8] A particular concern in this regard is the spread of dis- and misinformation about Covid-19 on social media sites, which has led the WHO to host a page with “myth busters” on their website and engage in discussions with social media companies.[9] Understanding what the general public knows about Covid-19 and which misperceptions they hold about the condition is important for US and UK public health authorities as well as the media to design effective information campaigns.

The speed with which Covid-19 is spreading across the world calls for rapid assessments of the population’s knowledge and perceptions of this infection.[7] This study tests a rapid online survey methodology to determine knowledge and misperceptions of Covid-19 among the general adult population in both the US and the UK.

## Methods

### Sampling participants

This study is a cross-sectional online survey conducted on the research platform created and managed by Prolific Academic Ltd.. Prolific is an online platform that connects researchers with individuals around the world who are interested in participating in online research studies.[10] The platform’s pool of participants numbers approximately 80,000 individuals of whom circa 43% reside in the UK and 33% in the US.[11] Researchers are required to pay participants a minimum of $6.50 per hour.

For this study, Prolific established strata by age group (18 - 27, 28 - 37, 38 - 47, 48 - 57, or ≥58 years), sex (male or female), and ethnicity (White, Black or African American, Asian or Asian Indian, Mixed, or “Other”), as well as all combinations of these strata. Using numbers from the latest census in the US and the UK, a given number of places for taking the questionnaire was opened on the Prolific platform in each stratum to achieve the same distribution of participants by age, sex, and ethnicity as in the general population. The targeted total sample size in each country was 1,500. Participants’ eligibility for the open places in a particular stratum was determined based on the information they had entered in their profile when registering with Prolific. Eligible participants enrolled in the study on a first-come, first-served basis. The study was implemented in two rounds of 1,500 participants each in the US and the UK, such that the total target sample size in each country was 3,000. Participants had to have indicated that they are fluent in English when registering with Prolific to be eligible for this study.

### Data collection

The data were collected using the online questionnaire shown in **Text S1** and **Text S2**. Participants received US$1.50 (UK participants received the equivalent in pound sterling) for completing the questionnaire. Following an informed consent form, the questionnaire asked participants about i) the cause, current state, and future development of the Covid-19 epidemic; ii) the risk of a fatal disease course; iii) knowledge of symptoms and recommended healthcare-seeking behavior; iv) measures to prevent a Covid-19 infection; and v) their perception of the risk posed by individuals of East-Asian ethnicity in their community. In order to investigate to what degree dis- and misinformation about Covid-19 has affected the general public’s beliefs about the condition, participants were directly asked whether or not they believed several falsehoods listed on the WHO’s “myth busters” website,[12] which the WHO selected because they had been circulating on social media.[13] Specifically, the questionnaire asked whether receiving a letter or package from China poses a risk of infection, and whether using hand dryers, rinsing your nose with saline, eating garlic, applying sesame oil to the skin, taking antibiotics, and vaccinating against pneumonia are effective in preventing a SARS-CoV-2 infection. The questionnaire was built using Qualtrics software. Participants had to answer a question to reach the next question. Numerical entry questions did not allow for non-sensical inputs (e.g., percentage questions were restricted to inputs between 0 and 100).

### Data analysis

For binary and categorical response options, I computed the percentage of participants who selected each response. For binomial proportions, I calculated two-sided 95% confidence intervals using the Wilson score interval.[14] I did not use sampling weights given that this was not a probabilistic sample of adults and that the survey was already self-weighting by the age, sex, and ethnicity groups used to establish the strata for sampling.

Three types of data quality checks were performed. First, participants who took less than two minutes to complete the questionnaire were excluded from the analysis because I judged this to indicate random clicking. This resulted in the exclusion of two participants. Second, if some respondents used random clicking to obtain the $1.50 reward as fast as possible, then a bimodal distribution in the time taken to complete the survey might be expected (with one study population clicking as fast as possible and one reading the questions). I, therefore, plotted a histogram of the time taken to complete the survey. Third, participants were asked at the end of the questionnaire whether they looked up any answers online (*“It is natural to be tempted to look up the answer to a question, especially when it’s only a click away. For approximately how many of the questions did you first look up the answer on Google or somewhere else before responding? The answer to this question will not affect your payment in any way*.*”*) and if so, for which question. Those who self-reported looking up an answer online for a question were excluded from the analysis for that particular question. This was the case for 81 US participants and 63 UK participants who reported looking up the answer online on a median of 1 (IQR: 1 – 5) and 1 (IQR: 1 – 2.5) questions, respectively.

## Results

### Data collection time

Figure 1 shows that each of the two rounds of the survey took two to three days to conduct. There was no evidence of a bimodal distribution in the time taken to complete the survey (**Figure S1**).

**Figure 1.**
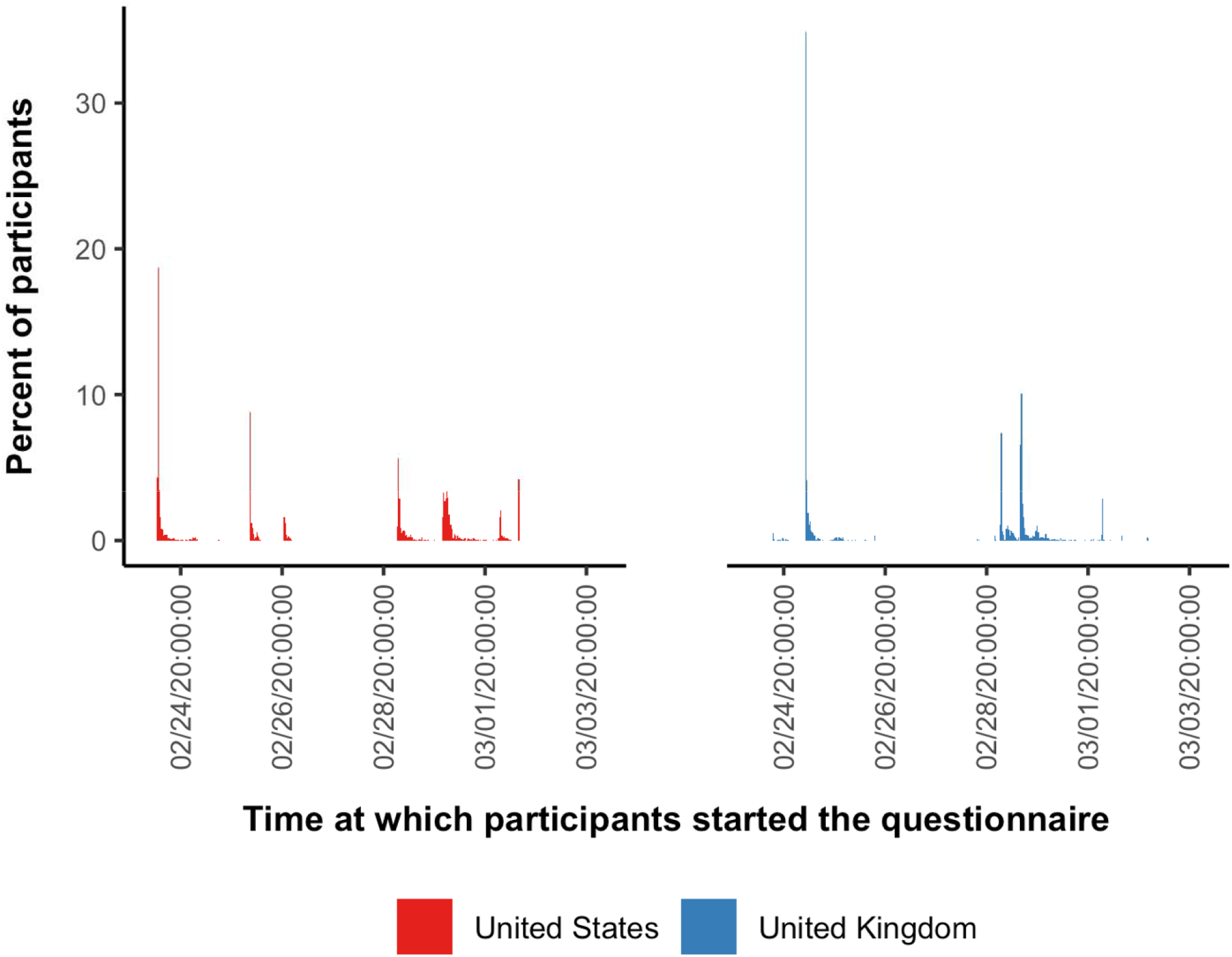
Time at which participants started the questionnaire.^1,2^ ^1^ Dates and times are in Pacific Standard Time. ^2^ Bins have a width equal to 30 minutes.

### Characteristics of the sample of adults who chose to participate in the study

Out of 3,000 adults residing in the US and 3,000 adults residing in the UK who could participate, 2,986 and 2,988, respectively, completed the questionnaire. Approximately two thirds (64.4% [1,924/2,986]) of US and half (51.5% [1,540/2,988]) of UK participants had a tertiary education degree (**Table 1**). 67.5% (2,015/2,986) of US participants had a total household income between $20,000 and $99,999, and 74.4% (2,223/2,988) of UK participants had a total household income between £15,000 and £74,999. 17.3% (516/2,986) of US participants and 13.7% (409/2,988) of UK participants were current students.

**Table 1.** Sample characteristics.

|  | United States | United Kingdom |
| --- | --- | --- |
| *n* | 2,986 | 2,988 |
| Female, *n* (%) | 1,519 (50.9) | 1531 (51.2) |
| Age group, *n* (%) |  |  |
| 18 – 27 years | 655 (21.9) | 550 (18.4) |
| 28 – 37 years | 687 (23.0) | 557 (18.6) |
| 38 – 47 years | 531 (17.8) | 563 (18.8) |
| 48 – 57 years | 493 (16.5) | 480 (16.1) |
| ≥ 58 years | 620 (20.8) | 838 (28.0) |
| Education, *n* (%) |  |  |
| Less than a high school diploma / A-Levels | 24 (0.8) | 396 (13.3) |
| High school degree / Completed A-Levels | 334 (11.2) | 682 (22.8) |
| Some undergraduate education (no degree) | 704 (23.6) | 370 (12.4) |
| Associate degree | 322 (10.8) | n.a.<sup>1</sup> |
| Bachelor's degree | 1068 (35.8) | 1030 (34.5) |
| Master's degree | 405 (13.6) | 330 (11.0) |
| Professional degree | 63 (2.1) | 100 (3.3) |
| Doctorate | 66 (2.2) | 80 (2.7) |
| Total household income, *n* (%) |  |  |
| <\$10,000 / <£7,500 | 165 (5.5) | 172 (5.8) |
| \$10,000 – \$19,000 / £7,500 - £14,999 | 222 (7.4) | 333 (11.1) |
| \$20,000 – \$29,000 / £15,000 - £22,499 | 342 (11.5) | 463 (15.5) |
| \$30,000 – \$39,000 / £22,500 - £29,999 | 325 (10.9) | 473 (15.8) |
| \$40,000 – \$49,000 / £30,000 - £37,499 | 280 (9.4) | 358 (12.0) |
| \$50,000 – \$59,000 / £37,500 - £44,999 | 304 (10.2) | 312 (10.4) |
| \$60,000 – \$69,000 / £45,000 - £52,499 | 230 (7.7) | 242 (8.1) |
| \$70,000 – \$79,000 / £52,500 - £59,999 | 242 (8.1) | 156 (5.2) |
| \$80,000 – \$89,000 / £60,000 - £67,499 | 138 (4.6) | 121 (4.0) |
| \$90,000 – \$99,000 / £67,500 - £74,999 | 154 (5.2) | 98 (3.3) |
| \$100,000 – \$149,000 / £75,000 - £99,999 | 401 (13.4) | 168 (5.6) |
| ≥\$150,000 / ≥£100,000 | 183 (6.1) | 92 (3.1) |
| Race or ethnicity, *n* (%) |  |  |
| White | 2269 (76.0) | 2540 (85.0) |
| Black or African American | 392 (13.1) | 110 (3.7) |
| Asian or Asian Indian | 191 (6.4) | 227 (7.6) |
| Mixed | 74 (2.5) | 62 (2.1) |
| Other | 60 (2.0) | 49 (1.6) |
| Current student, *n* (%) | 516 (17.3) | 409 (13.7) |
| Chinese descent, *n* (%) |  |  |
| Born in China | 11 (0.4) | 15 (0.5) |
| Parents or grandparents born in China | 57 (1.9) | 27 (0.9) |
| Works as a healthcare provider, n (%) |  |  |
| Nurse | 33 (1.1) | 44 (1.5) |
| Physician | 5 (0.2) | 15 (0.5) |
| Pharmacist | 10 (0.3) | 6 (0.2) |
| Other | 102 (3.4) | 118 (3.9) |

<sup>†</sup> Associate degrees are not awarded in the UK.

### Cause, current state, and future development of the Covid-19 epidemic

On a seven-point Likert scale ranging from ‘extremely unlikely’ to ‘extremely likely’, 23.9% (22.4% – 25.5%) of US participants and 18.4% (17.1% – 19.9%) of UK participants selected ‘slightly likely’, ‘moderately likely’, or ‘extremely likely’ when asked whether Covid-19 is a bioweapon developed by a government or terrorist organization (**Figure S2**). US and UK participants estimated that a median of 100 (IQR: 20 – 500) and 40 (IQR: 13 – 200) individuals in their country, respectively, were currently infected with Covid-19. 61.0% (59.3% – 62.8%) of US and 71.7% (70.1% – 73.3%) of UK participants thought that the number of fatalities from Covid-19 in their country will be 500 people or less by the end of 2020 (**Figure 2**).

**Figure 2.**
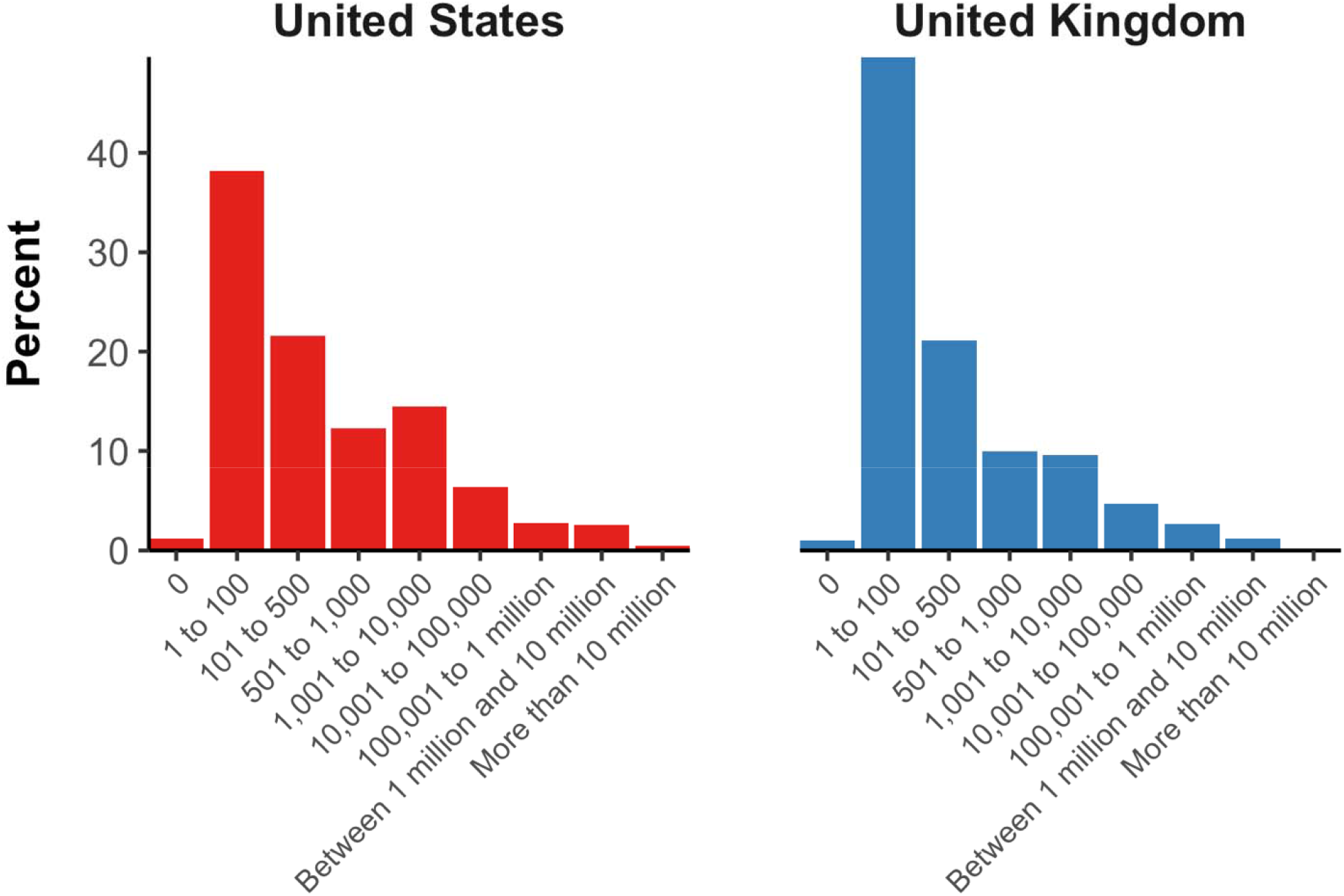
Percent of participants who selected each category for their estimate of the number of Covid-19 deaths in their country by the end of 2020.

### Case fatality rate

When asked what percent of individuals infected with Covid-19 experience a fatal disease course, the median estimate given by participants was 5.0% (IQR: 2.0% – 15.0%) among US participants and 3.0% (IQR: 2.0% – 10.0%) among UK participants. The full distribution of responses, as well as a magnification of the distribution of responses among those who estimated a risk of death ≤10%, is shown in **Figure 3**.

**Figure 3.**
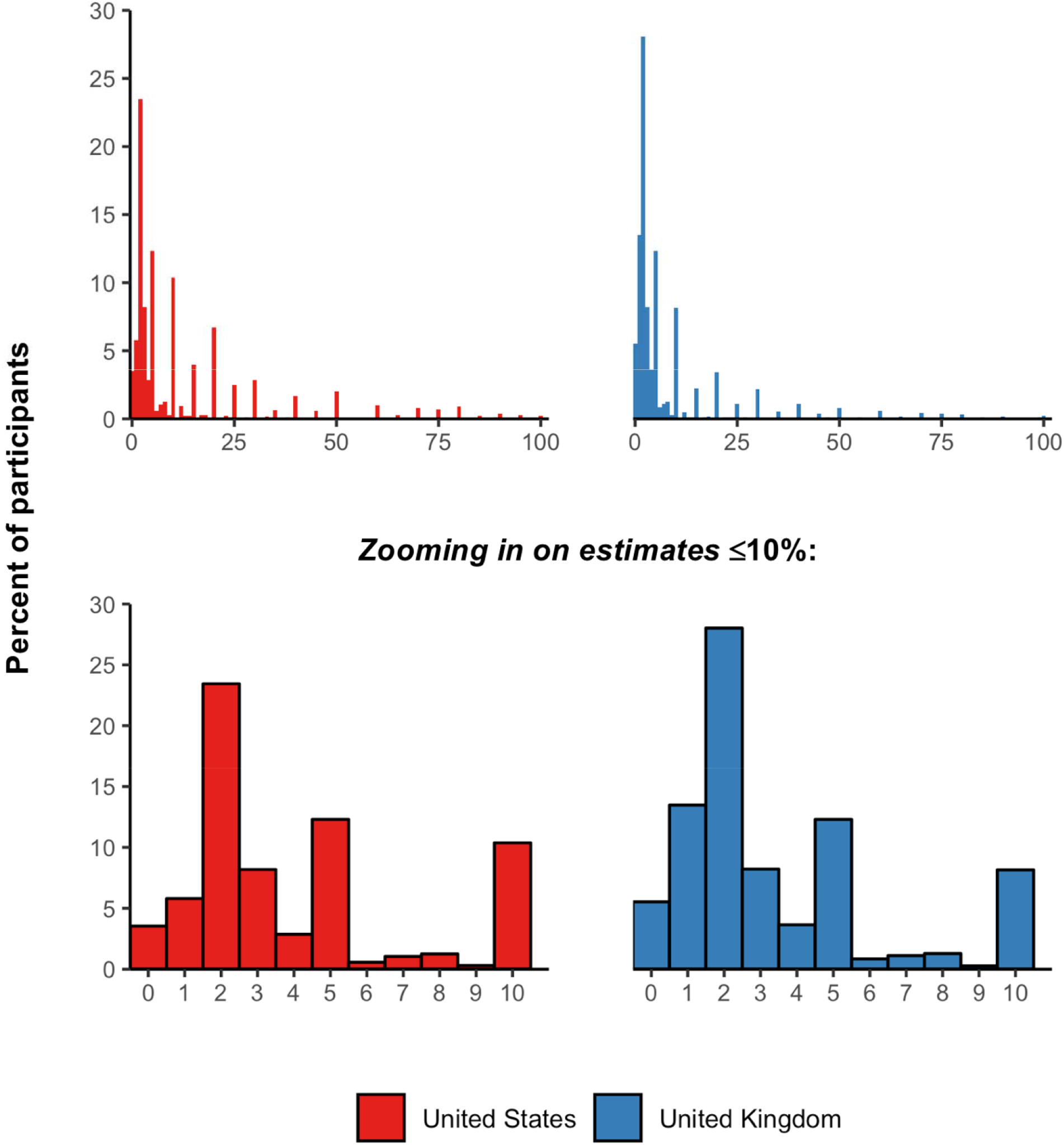
Distribution of responses to the question “What percent of people who get infected with the new coronavirus die from this infection?”

When asked “when they have been infected, what age groups are most likely to die from the illness caused by the new coronavirus” and presented with the option to select ‘children’, ‘young adults’, or ‘older adults’ (selecting more than option was possible), 96.3% (95% CI: 95.6% – 96.9%) of participants in the US and 98.3% (95% CI: 97.7% – 98.7%) of participants in the UK selected ‘older adults’. However, 53.8% (95% CI: 52.1% – 55.6%) and 39.1% (95% CI: 37.4% – 40.9%) of participants in the US and the UK, respectively, also thought that children were at a high risk of death when infected. Almost all participants in both countries (96.3% [95% CI: 95.6% – 97.0%] in the US and 97.5% [95% CI: 96.9% – 98.0%] in the UK) responded that adults with other health problems were more likely to experience a fatal disease course than those without any other health problems.

### Symptoms of COVID-19 and recommended healthcare-seeking behavior

Most participants in both the US and the UK recognized fever, cough, and shortness of breath as three common symptoms and signs of a Covid-19 infection (**Figure 4**).

**Figure 4.**
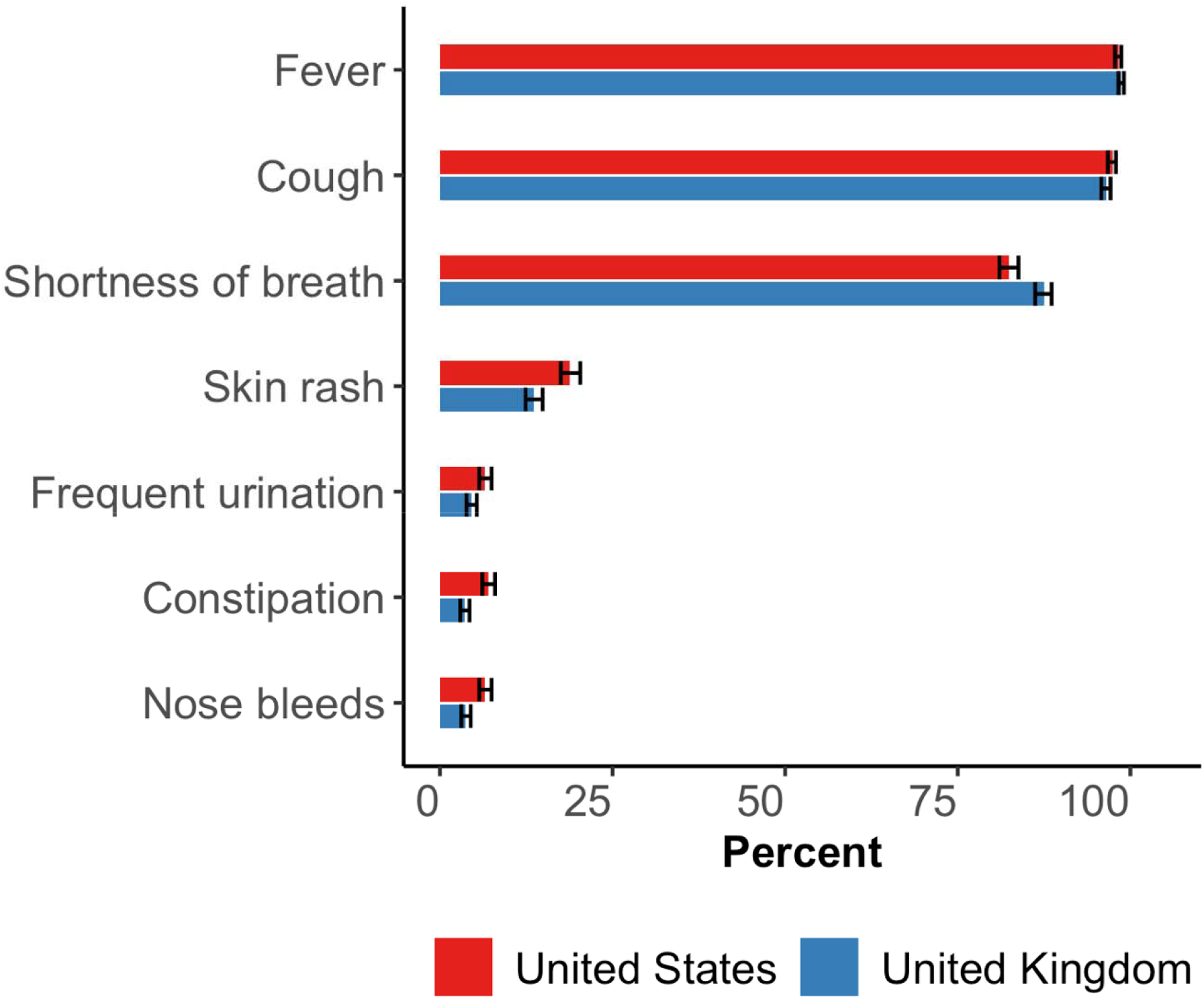
Percent of participants who replied with “yes” to whether each of seven symptoms or signs were common in a Covid-19 infection^1^ ^1^ The horizontal black bars depict 95% confidence intervals using Wilson’s method.[14]

When asked *“if you have a fever or cough and recently visited China, or spent time with someone who did, what would be the best course of action?”*, 64.2% (95% CI: 62.4% – 65.9%) of US participants and 79.0% (95% CI: 77.5% – 80.5%) of UK participants responded with the recommended care-seeking option of staying home and contacting their health system. About a third of respondents stated that they would either delay care-seeking, attend the hospital emergency department unannounced, or take a taxi or public transport to their primary care provider (**Figure 5**).

**Figure 5.**
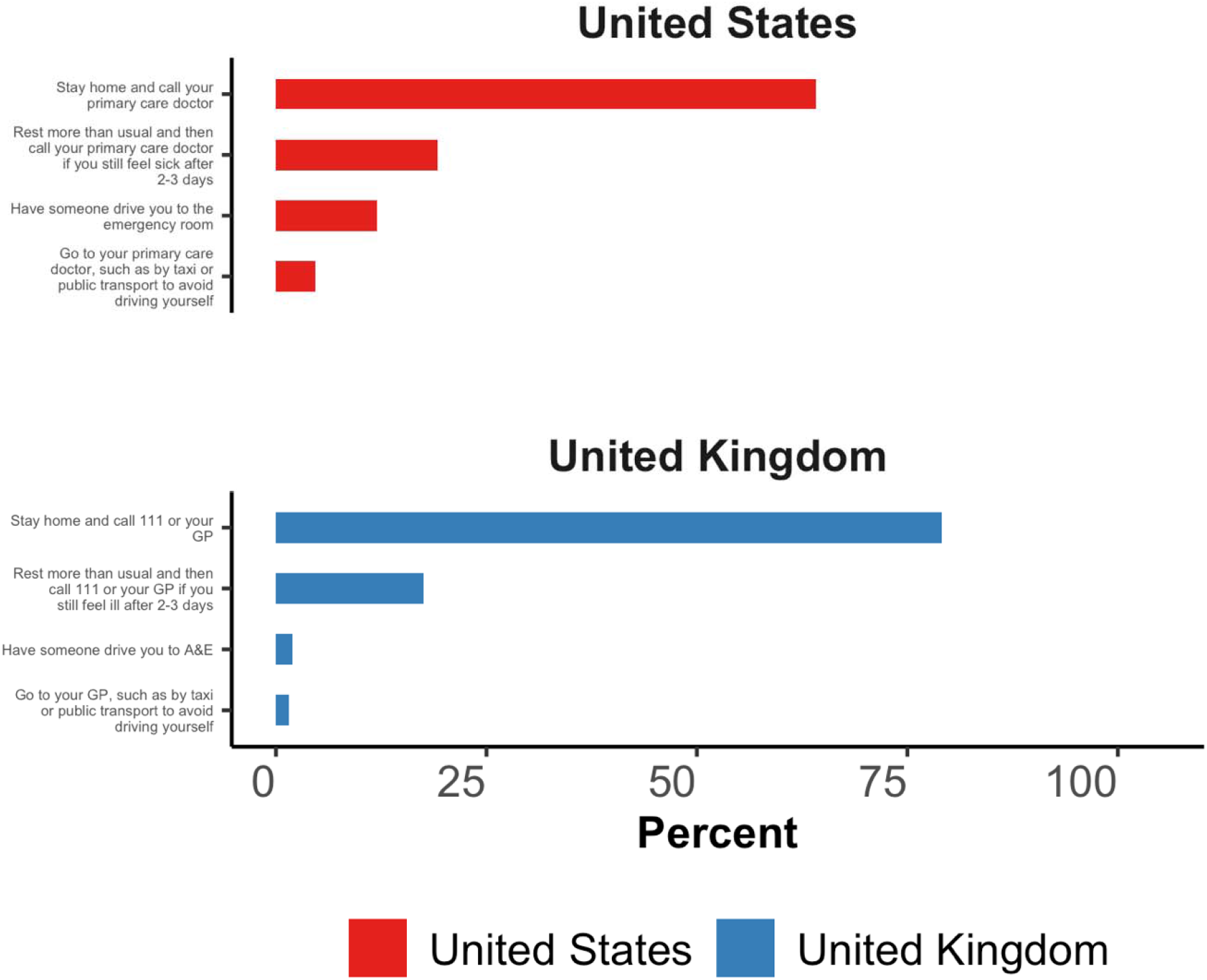
Responses to the question “If you have a fever or cough and recently visited China, or spent time with someone who did, what would be the best course of action?”

### Preventing a Covid-19 infection

92.6% (95% CI: 91.6% – 93.4%) of US and 86.0% (95% CI: 84.7% – 87.2%) of UK participants selected each of the following three responses as being effective in preventing infection with SARS-CoV-2: washing your hands, avoiding close contact with people who are sick, and avoiding touching your eyes, nose, and mouth with unwashed hands (**Figure 6**). However, a substantial proportion of participants also thought that using a hand dryer, rinsing your nose with saline, taking antibiotics, and gargling mouthwash were effective prevention measures. 43.5% (95% CI: 41.7% – 45.2%) and 36.0% (95% CI: 34.3% – 37.8%) of US and UK participants, respectively, selected at least one of these options. 37.8% (95% CI: 36.1% – 39.6%) of US participants and 29.7% (95% CI: 28.1% – 31.4%) of UK participants agreed with the following statement: *“Consistently wearing a face mask is highly effective in protecting you from getting infected with the new coronavirus. For the purpose of this question, “highly effective” is defined as reducing your risk of getting infected by >95% and a “face mask” is a common medical mask*.*”*

**Figure 6.**
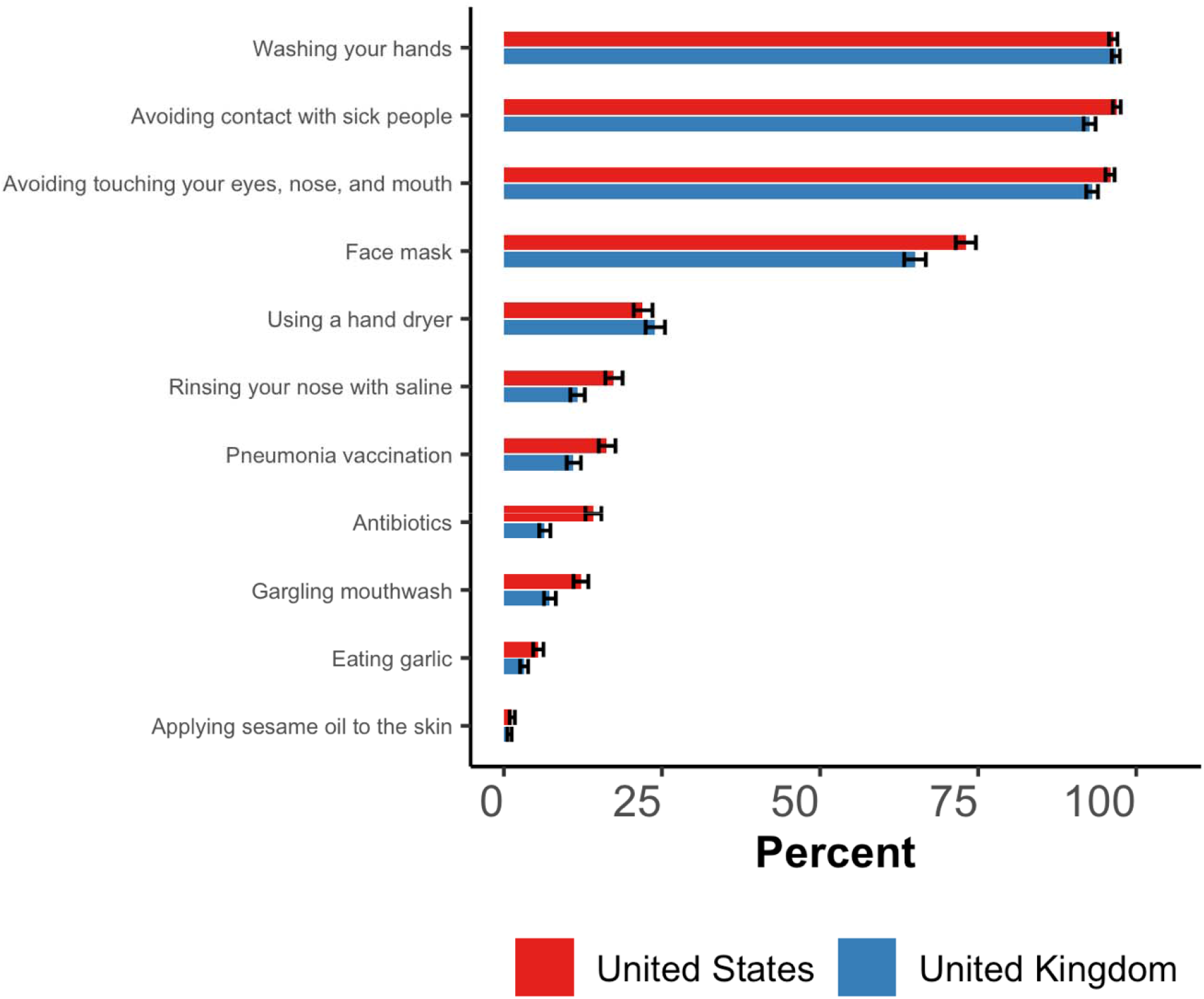
Percent of participants who replied with ‘yes’ to whether each of eleven actions help prevent an infection with Covid-19.^1^ ^1^ The horizontal black bars depict 95% confidence intervals using Wilson’s method.[14]

74.8% (95% CI: 73.2% – 76.4%) of US participants and 81.2% (95% CI: 79.8% – 82.6%) of UK participants correctly selected *“droplets of saliva that land in the mouths or noses of people who are nearby when an infected person sneezes or coughs”* as the main mode of Covid-19 transmission (**Figure S3**). Virtually all participants disagreed with the statement that *“only older adults can become infected with the new coronavirus”* (96.5% [95% CI: 95.8% – 97.1%] of US participants and 97.1% [95% CI: 96.5 – 97.7%] of UK participants) and thought that there is currently no vaccine available that protects against Covid-19 (96.0% [95% CI: 95.3% – 96.7%] of US participants and 97.5% [95% CI: 96.9% – 98.0%] of UK participants). More than 20% of participants in both the US and the UK thought that their government should quarantine everyone coming in from abroad for 14 days and suspend all air travel to their country (**Figure 7**).

**Figure 7.**
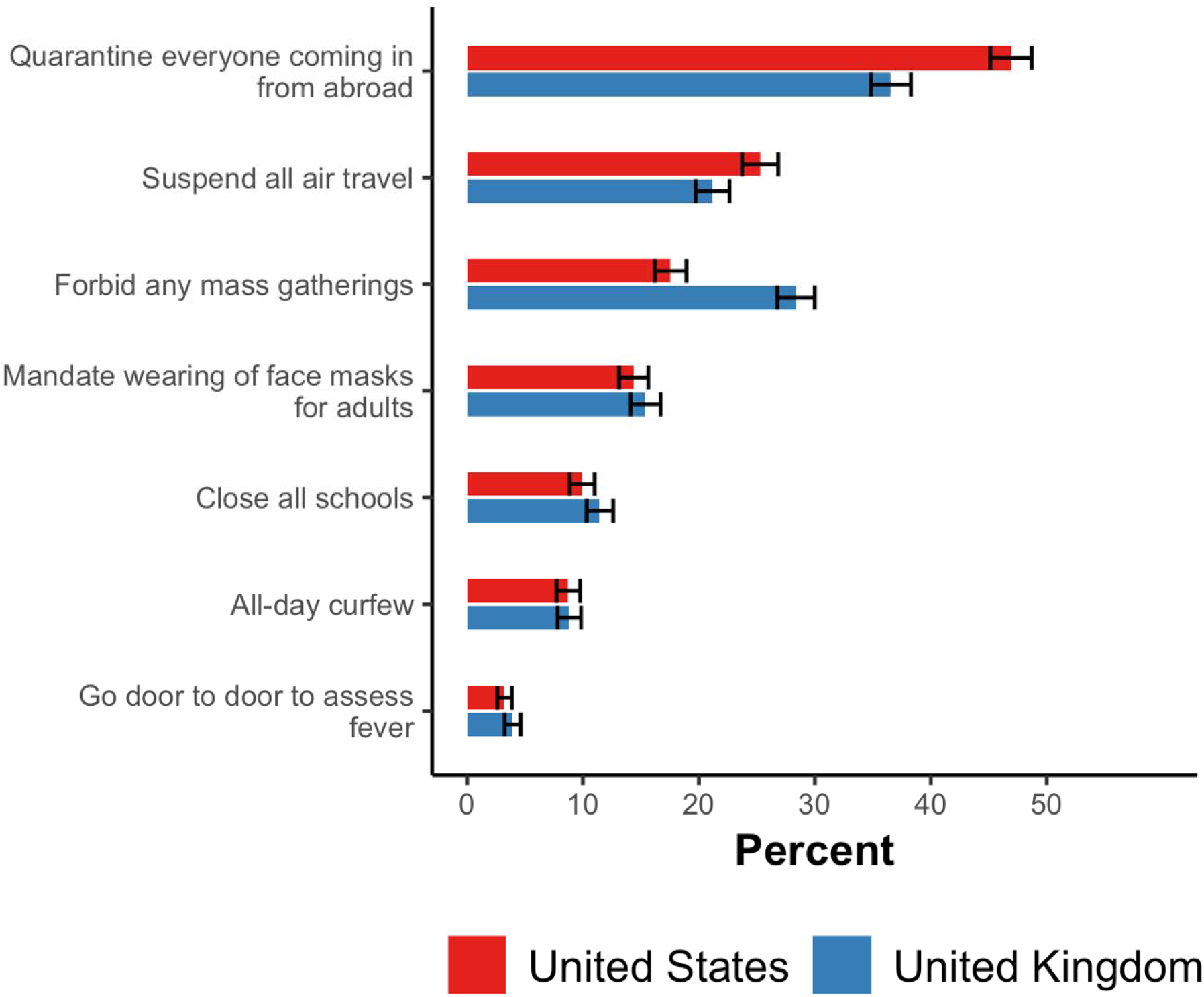
Percent of participants who replied with ‘yes’ to each government action in response the question “At this point in the coronavirus epidemic, do you think your government should implement the following measures to prevent spreading of the virus?”.^1^ ^1^ The horizontal black bars depict 95% confidence intervals using Wilson’s method.[14]

### Perceptions of the risk posed by community members of East-Asian ethnicity

When asked about the prevalence of an infection with SARS-CoV-2 among East-Asian individuals in their country, the median estimate among US and UK participants was 0.5% (IQR: 0.0% – 2.0%) and 0.5% (IQR: 0.0% – 1.0%), respectively (**Figure 8**). The median rose to 1.0% (IQR: 0.0% – 5.0%) among both US and UK participants when asking about the prevalence of Covid-19 among *“adults of East-Asian ethnicity in your neighborhood who wear a face mask”*. 25.6% (95% CI: 24.1% – 27.2%) of US participants and 29.6% (95% CI: 28.0% – 31.3%) of UK participants responded with ‘yes’ to the question *“Do you think it would be prudent for you to not eat at Chinese restaurants for the next few weeks to reduce the risk of getting infected with the new coronavirus?”*. Approximately a quarter of participants (29.0% [27.4%– 30.7%] of US participants and 24.4% [22.9% – 26.0%] of UK participants) thought one may become infected with Covid-19 by receiving a package from China. When asked *“If you were an Uber driver today, would you try to reject ride requests from people with East Asian-sounding names (or a profile photo of East-Asian ethnicity) to reduce your risk of getting infected with the new coronavirus?”*, 29.7% (95% CI: 28.1% – 31.3%) of US participants and 40.8% (95% CI: 39.0% – 42.5%) of UK participants responded with ‘sometimes’, ‘often’, or ‘always’ (**Figure 9**).

**Figure 8.**
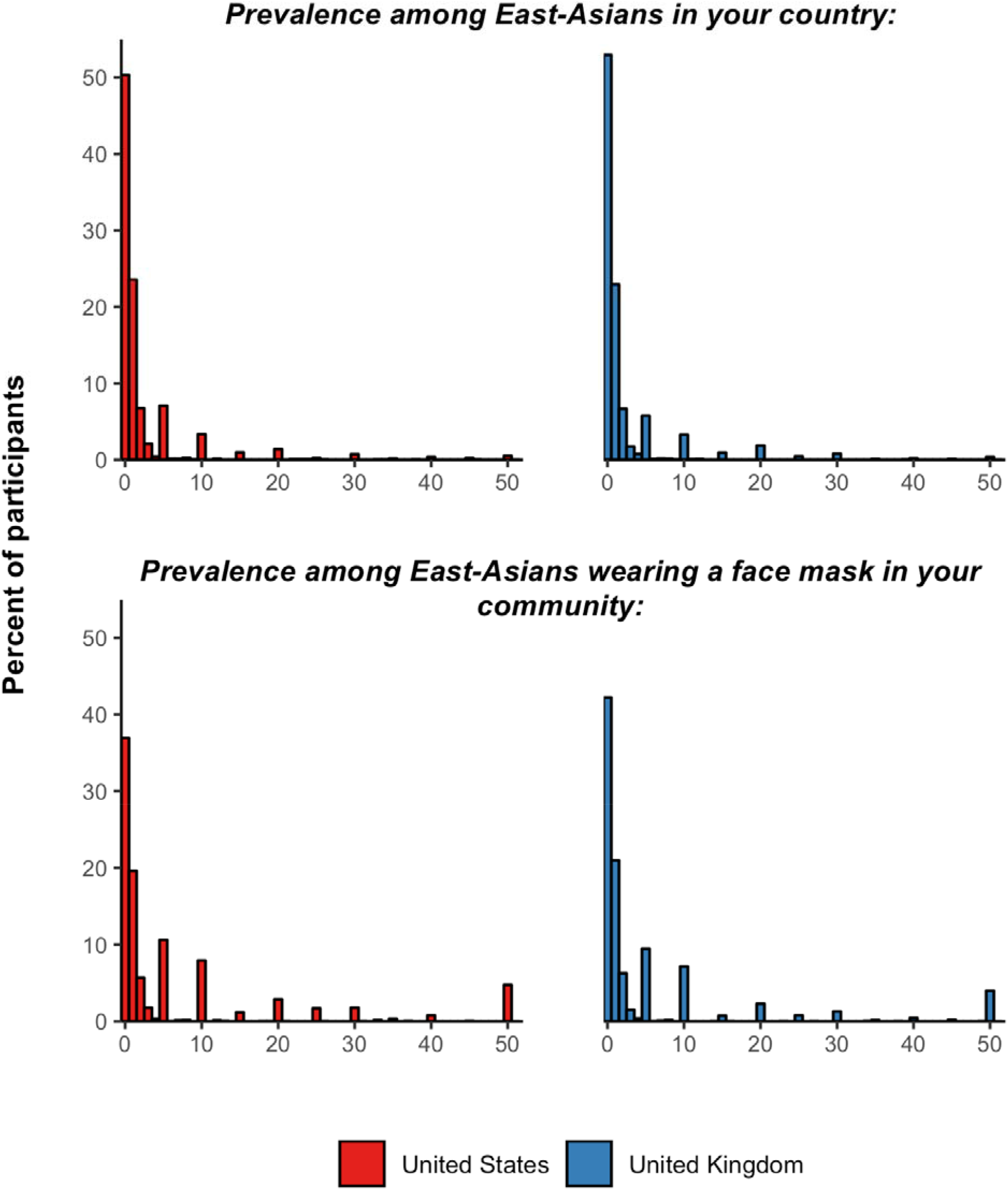
Distribution of the responses to questions on Covid-19 prevalence among individuals of East-Asian ethnicity^1^ ^1^ 32 and 129 participants estimated a prevalence greater than 50% for the prevalence among East-Asian individuals in their country and East-Asian individuals wearing a face mask in their community, respectively. The responses from these individuals are not shown in the histogram above.

**Figure 9.**
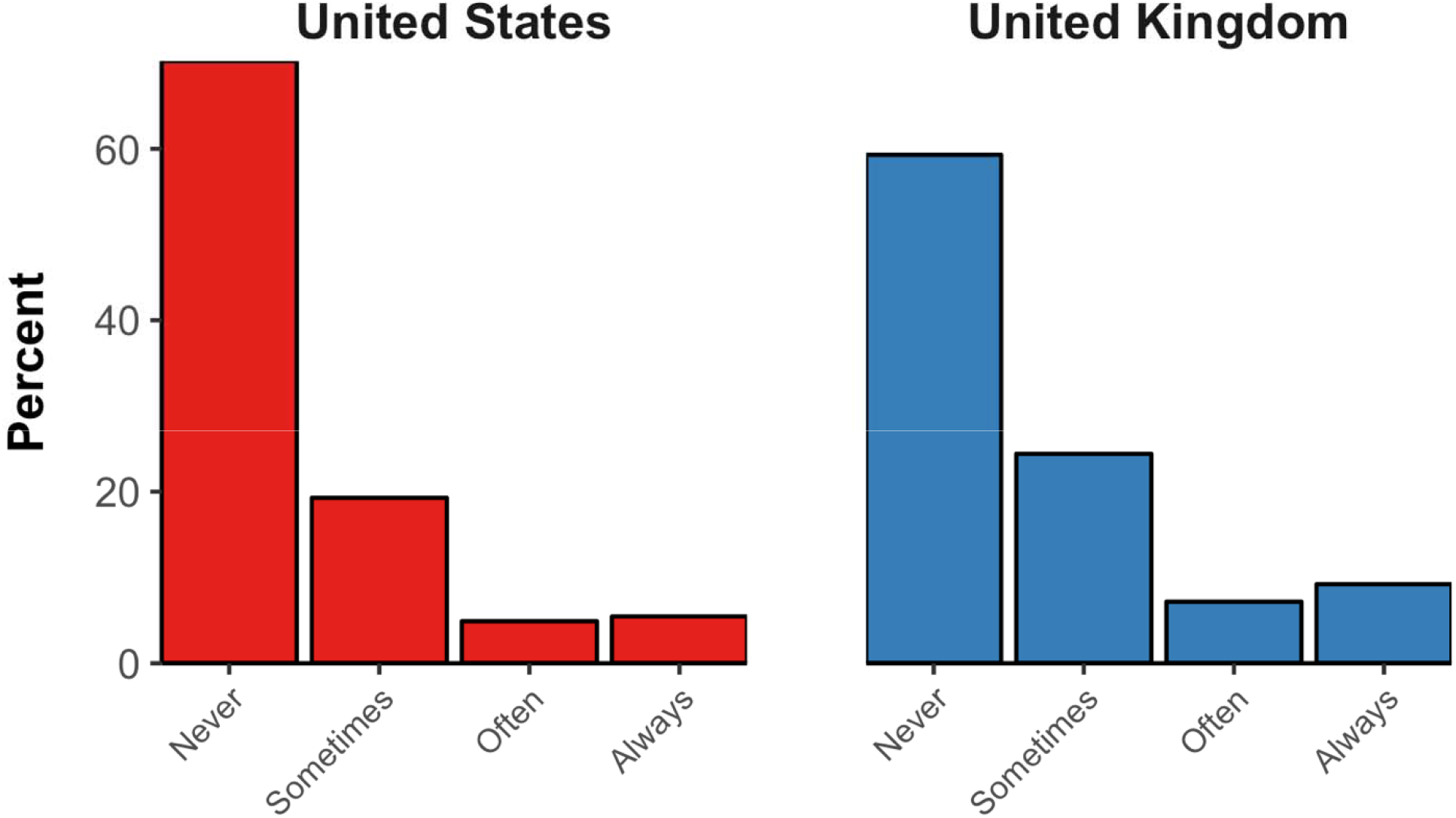
Distribution of responses to the question “If you were an Uber driver today, would you try to reject ride requests from people with East Asian-sounding names (or a profile photo of East-Asian ethnicity) to reduce your risk of getting infected with the new coronavirus?”

## Discussion

It was possible to conduct an in-depth online assessment of knowledge and perceptions of Covid-19 among the general public in the US and the UK in a short time frame. It took two to three days to obtain a completed questionnaire of 22 knowledge and perception questions from 1,500 adults in each the US and the UK, when allowing enrolment only in relatively granular strata by age, sex, and ethnicity (and each of these variables’ combinations). Importantly, the distribution of participants by education and household income in this sample, even though not part of the enrolment criteria, was similar to that of the general population in the US and the UK.[15, 16] In terms of data quality, there was no indication that participants randomly clicked on responses to earn the monetary reward as quickly as possible, with only two participants having taken less than two minutes to complete the questionnaire and there being no evidence of a bimodal distribution in the time taken to complete the questionnaire. The direct cost to Prolific of administering the questionnaire was merely $8,961 ($1.50 per completed questionnaire for a total of 5,974 participants).

Regarding the survey findings, the general public in both the US and the UK held several important misconceptions about Covid-19. Participants in both countries generally overestimated the probability of a fatal disease course among those infected with Covid-19 (the case fatality rate is currently believed to be between one and two percent among reported cases,[5, 17, 18] but could be substantially lower if there are many unreported and/or asymptomatic cases[19]), thought that children were at an especially high risk of death from Covid-19 (which is currently not believed to be the case[5, 18-20]), and believed that common surgical masks are highly effective in protecting them from catching a Covid-19 infection. Participants also likely overestimated the prevalence of Covid-19 among East-Asian individuals in their communities. Relatedly, a substantial proportion thought that they should refrain from frequenting Chinese restaurants, stated that they would refuse Uber rides to individuals of East-Asian ethnicity, and perceived that receiving a package from China poses a risk of a Covid-19 infection. In general, differences in knowledge and misperceptions between US and UK participants were small.

This study’s findings on levels of knowledge and prevalence of misconceptions regarding Covid-19 could inform relevant information campaigns by public health authorities and the media, as well as communication of healthcare workers with patients. For instance, such information provision may highlight the comparatively low case fatality rate, the low risk posed by individuals of East-Asian ethnicity living in the US and the UK, and that children do not appear to be at a heightened risk of dying from Covid-19. In addition, a substantial proportion of participants appeared to believe that common surgical masks are highly effective in preventing infection with SAES-CoV-2. Information campaigns may, therefore, want to emphasize the comparative effectiveness of common surgical masks versus other methods of prevention, particularly frequent and thorough handwashing and avoiding close contact with people who are sick. Lastly, it is important to note that while the general public appeared to be well informed regarding the common symptoms of Covid-19, over a fourth of participants selected a healthcare-seeking option that could lead to further transmission of Covid-19. Thus, clear messaging on the recommended care-seeking action when experiencing some of the core symptoms of Covid-19 will be crucial.

Public health information campaigns may also want to directly target some of the mis- and disinformation that have circulated on social media.[9, 12, 13, 21] Such measures could include information that rinsing your nose with saline, using a hand dryer, taking antibiotics, and gargling mouthwash are not effective prevention measures, and that receiving a letter or package from China does not pose a risk of Covid-19 infection. These are all falsehoods listed on the WHO’s “myth busters” website that a substantial proportion of participants in this study believed.[12] More broadly, this study underscores the need for the WHO and other public health bodies to continue working with social media campaigns to minimize the circulation of inaccurate information about Covid-19. In line with recent media reporting that this conspiracy theory has been actively spread on Twitter,[21] about one in five participants believed it to be ‘slightly likely’, ‘moderately likely’, or ‘extremely likely’ that SARS-CoV-2 is a bioweapon developed by a government or terrorist organization.

Participants did not expect that a large number of individuals would die from Covid-19 in their country by the end of 2020. This finding may be surprising considering that fear-inducing headlines in the media may (at least up to a certain extent[22]) result in more attention by readers than more emotionally neutral ones, which could result in a catastrophizing of the epidemic. Moving forward, information campaigns on Covid-19 may need to balance the messaging of two important facts about the Covid-19 epidemic that could be interpreted by the general public to stand in direct conflict with each other. That is, on the one hand, the case fatality rate of Covid-19 appears to be lower compared to other recent infectious disease outbreaks, such as Ebola,[23] the severe acute respiratory syndrome (SARS),[24] and the Middle East respiratory syndrome (MERS).[24, 25] On the other hand, however, the epidemic could cause a large number of fatalities, which implies that actions by governments and the general public to reduce transmission of Covid-19 could save many lives.

This study has several limitations. First, while the sample of participants is representative of the US and UK general population by age, sex, and ethnicity, and the distribution of participants by household income and education was similar to that in the US and UK general population, participants may still differ from the general population on a variety of other characteristics. These characteristics may be both correlated with their knowledge and perceptions of Covid-19 as well as with their decision to participate in the study and/or to create a profile with Prolific. Second, the estimates of discrimination against individuals of East-Asian ethnicity may be an underestimate because some participants may not have wanted to volunteer their discriminating tendencies to themselves or to the researcher. However, I as the researcher had no access to any identifying information about the research participants and participants were reminded of this fact prior to answering the question. In addition, such social desirability bias has been found to be lower in online surveys than in telephone or in-person surveys.[26, 27] Third, it was possible for participants to randomly click responses in order to devote the least amount of time to earn the $1.50 reward. In my view, this issue is unlikely to have caused major bias in this study because i) there was no evidence of a bimodal distribution in the time taken to complete the survey (see Figure S1), ii) while it was physically possible to complete the survey in under 90 seconds when randomly clicking on responses, only two participants completed the survey in under two minutes, and iii) the monetary reward ($1.50) was relatively small and thus, for most participants, unlikely to be the main motivation for participating in the study. Lastly, it is possible that individuals looked up the answers to some of the questions online prior to answering, which may have biased the results from factual (rather than opinion-focused) questions. Participants were reminded of the importance not to look up answers online prior to taking the survey and were asked at the end of the survey (while being reassured that their payment is not influenced by whether they volunteer information on having looked up an answer online) for which, if any, questions they searched for an answer online prior to responding.

Rapid online surveys are a promising method to assess and track knowledge and perceptions in the midst of rapidly evolving infectious disease outbreaks. Such assessments are crucial because ensuring that the general public is well informed about a condition like Covid-19 could reduce unnecessary anxiety as well as reduce disease transmission and, thus, ultimately save lives.

## Data Availability

All data and statistical code for this study is available at: https://purl.stanford.edu/tr461wp6422

## Acknowledgements

PG conceived of the study, designed the questionnaire, conducted the data analysis, and wrote the manuscript. PG is the guarantor of the work. PG was supported by the National Center for Advancing Translational Sciences of the National Institutes of Health under Award Number KL2TR003143. The funder had no role in study design, data collection, data analysis, data interpretation, or writing of the report. PG had full access to all the data in the study and had final responsibility for the decision to submit for publication.

I would like to thank all participants for their time and effort, and the team at Prolific Academic Ltd. for publishing this study to their pool of research participants.

## Conflicts of Interest

None declared.

## Notes

### Competing Interest Statement

The authors have declared no competing interest.

## References

1. Gong W, Taighoon Shah M, Firdous S, Jarrett BA, Moulton LH, Moss WJ, et al. Comparison of three rapid household survey sampling methods for vaccination coverage assessment in a peri-urban setting in Pakistan. Int J Epidemiol. 2019 Apr 1;48(2):583-95. PMID: 30508112. doi: 10.1093/ije/dyy263.

2. Kennedy C, Hartig H. Response rates in telephone surveys have resumed their decline. Washington, D.C., USA: Pew Research Center; 2019 [cited 2020 March 1]; Available from: https://www.pewresearch.org/fact-tank/2019/02/27/response-rates-in-telephone-surveys-have-resumed-their-decline/.

3. Katharine G. Abraham, Sara Helms, Stanley Presser. How Social Processes Distort Measurement: The Impact of Survey Nonresponse on Estimates of Volunteer Work in the United States. American Journal of Sociology. 2009;114(4):1129–65. doi: 10.1086/595945.

4. Charles QL, Alexandra C, Leenisha M, Ashley A. In Search of the Optimal Mode for Mobile Phone Surveys in Developing Countries. A Comparison of IVR, SMS, and CATI in Nigeria. Survey Research Methods. 2019 12/10;13(3). doi: 10.18148/srm/2019.v13i3.7375.

5. WHO-China Joint Mission on Coronavirus Disease 2019. Report of the WHO-China Joint Mission on Coronavirus Disease 2019 (COVID-19). Geneva, Switzerland: World Health Organization, 2020.

6. World Health Organization. WHO Director-General’s opening remarks at the media briefing on COVID-19 - 11 March 2020. Geneva, Switzerland: World Health Organization; 2020 [cited 2020 March 16 2020]; Available from: https://www.who.int/dg/speeches/detail/who-director-general-s-opening-remarks-at-the-media-briefing-on-covid-1911-march-2020.

7. Dong E, Du H, Gardner L. An interactive web-based dashboard to track COVID-19 in real time. Lancet Infect Dis. 2020 Feb 19. PMID: 32087114. doi: 10.1016/s1473-3099(20)30120-1.

8. Janz NK, Becker MH. The Health Belief Model: A Decade Later. Health Education Quarterly. 1984;11(1):1-47. PMID: 6392204. doi: 10.1177/109019818401100101.

9. Zarocostas J. How to fight an infodemic. The Lancet. 2020 2020/02/29/;395(10225):676. doi: https://doi.org/10.1016/S0140-6736(20)30461-X.

10. Prolific Academic Ltd. How it works. London, UK: Prolific Academic Ltd.; 2020 [cited 2020 February 29]; Available from: https://www.prolific.co/#researcher-content.

11. Prolific. Explore our participant pool demographics. London, UK: Prolific; 2020 [cited 2020 February 17]; Available from: https://www.prolific.co/demographics.

12. World Health Organization. Coronavirus disease (COVID-19) advice for the public: Myth busters. Geneva, Switzerland: World Health Organization; 2020 [cited 2020 February 29]; Available from: https://www.who.int/emergencies/diseases/novel-coronavirus-2019/advice-for-public/myth-busters.

13. Richtel M. W.H.O. Fights a Pandemic Besides Coronavirus: an ‘Infodemic’. The New York Times. 2020.

14. Wilson EB. Probable Inference, the Law of Succession, and Statistical Inference. Journal of the American Statistical Association. 1927;22(158):209–12. doi: 10.2307/2276774.

15. United States Census Bureau. Explore data. Washington, D.C.: United States Census Bureau; 2020 [cited 2020 March 1]; Available from: https://www.census.gov/data.html.

16. Office for National Statistics. People, population and community. London, UK: Office for National Statistics; 2020 [cited 2020 March 1]; Available from: https://www.ons.gov.uk/peoplepopulationandcommunity.

17. Wu Z, McGoogan JM. Characteristics of and Important Lessons From the Coronavirus Disease 2019 (COVID-19) Outbreak in China: Summary of a Report of 72314 Cases From the Chinese Center for Disease Control and Prevention. JAMA : the journal of the American Medical Association. 2020 Feb 24. PMID: 32091533. doi: 10.1001/jama.2020.2648.

18. Guan W-j, Ni Z-y, Hu Y, Liang W-h, Ou C-q, He J-x, et al. Clinical Characteristics of Coronavirus Disease 2019 in China. New England Journal of Medicine. 2020. doi: 10.1056/NEJMoa2002032.

19. Fauci AS, Lane HC, Redfield RR. Covid-19 — Navigating the Uncharted. New England Journal of Medicine. 2020. doi: 10.1056/NEJMe2002387.

20. Centers for Disease Control and Prevention. Frequently Asked Questions and Answers: Coronavirus Disease-2019 (COVID-19) and Children. Atlanta, GA, USA: Centers for Disease Control and Prevention,; 2020 [cited 2020 February 29]; Available from: https://www.cdc.gov/coronavirus/2019-ncov/specific-groups/children-faq.html.

21. Romm T. Millions of tweets peddled conspiracy theories about coronavirus in other countries, an unpublished U.S. report says. The Washington Post. 2020.

22. Kim HS, Forquer H, Rusko J, Hornik RC, Cappella JN. Selective Exposure to Health Information: The Role of Headline Features in the Choice of Health Newsletter Articles. Media Psychology. 2016 2016/10/01;19(4):614–37. doi: 10.1080/15213269.2015.1090907.

23. Jacob ST, Crozier I, Fischer WA, 2nd, Hewlett A, Kraft CS, Vega MA, et al. Ebola virus disease. Nature reviews Disease primers. 2020 Feb 20;6(1):13. PMID: 32080199. doi: 10.1038/s41572-020-0147-3.

24. de Wit E, van Doremalen N, Falzarano D, Munster VJ. SARS and MERS: recent insights into emerging coronaviruses. Nature reviews Microbiology. 2016 Aug;14(8):523-34. PMID: 27344959. doi: 10.1038/nrmicro.2016.81.

25. Mahase E. Coronavirus: covid-19 has killed more people than SARS and MERS combined, despite lower case fatality rate. BMJ. 2020;368:m641. doi: 10.1136/bmj.m641.

26. Kreuter F, Presser S, Tourangeau R. Social Desirability Bias in CATI, IVR, and Web Surveys: The Effects of Mode and Question Sensitivity. Public Opinion Quarterly. 2009;72(5):847–65. doi: 10.1093/poq/nfn063.

27. Keeter S. From Telephone to the Web: The Challenge of Mode of Interview Effects in Public Opinion Polls. Washington, D.C., USA: Pew Research Center; 2015 [cited 2020 March 1]; Available from: https://www.pewresearch.org/methods/2015/05/13/from-telephone-to-the-web-the-challenge-of-mode-of-interview-effects-in-public-opinion-polls/.

